# Assessing DxGPT: Diagnosing Rare Diseases with Various Large Language Models

**DOI:** 10.1101/2024.05.08.24307062

**Authors:** Juanjo do Olmo, Javier Logroño, Carlos Mascías, Marcelo Martínez, Julián Isla

## Abstract

Diagnosing rare diseases is a significant challenge in healthcare, with patients often experiencing long delays and misdiagnoses. The large number of rare diseases and the difficulty for doctors to be familiar with all of them contribute to this problem. Artificial intelligence, particularly large language models (LLMs), has shown promise in improving the diagnostic process by leveraging their extensive knowledge to help doctors navigate the complexities of diagnosing rare diseases.

Foundation 29 presents a comprehensive evaluation of DxGPT, a web-based platform designed to assist healthcare professionals in the diagnostic process for rare diseases. The platform currently utilizes GPT-4, but this study also compares its performance with other large language models, including Claude 3, Gemini 1.5 Pro, Llama, Mistral, Mixtral, and Cohere Command R+. It is crucial to emphasize that DxGPT is not a medical device but rather a decision support tool that aims to aid in clinical reasoning.

This study extends beyond initial synthetic patient cases, incorporating real-world data from the RAMEDIS and Peking Union Medical College Hospital (PUMCH) datasets. The analysis followed two main metrics: Strict Accuracy (P1), how often the first diagnostic suggestion agreed with the real diagnosis, and Top-5 Accuracy (P1 + P5), how often the right diagnosis was in the top five suggestions. The results show a complex picture of diagnostic accuracy, with performance varying significantly across models and datasets:

- On the synthetic dataset, closed models like GPT-4, Claude, and Gemini exhibited relatively high accuracy. Open models like Llama 3 and Mixtral performed reasonably well, though lagging behind.
- On the RAMEDIS rare disease cases, Claude 3 Opus model demonstrated 55% Strict Accuracy and 70% Top-5 Accuracy, outperforming other closed models. Open models like Llama 3 and Mixtral showed moderate accuracy.
- The PUMCH dataset proved challenging for all models, with the highest Strict Accuracy at 59.46% (GPT-4 Turbo 1106) and Top-5 Accuracy at 64.86%.

These findings demonstrate the potential of DxGPT and LLMs in improving diagnostic methods for rare diseases. However, they also emphasize the need for further validation, particularly in real-world clinical settings, and comparison with human expert diagnoses. Successful integration of AI into medical diagnostics will require collaboration between researchers, clinicians, and regulatory bodies to ensure safety, efficacy, and ethical deployment.

## 1 Introduction

### 1.1 Background

All patients, but specially those with rare diseases often face a prolonged and challenging path to diagnosis. According to a study by Benito-Lozano et al. [1], 56.4% of patients experienced a delay in the diagnosis of their rare diseases, with the mean time taken falling between 5 [2] to 6 years. The International Rare Diseases Research Consortium (IRDiRC) defines diagnostic delay as any period exceeding a year. This delay often leads to anxiety, frustration, impacted relationships, and substantial costs due to unnecessary consultations [3]. Moreover, the vast number of rare diseases, estimated between 5,000 and 8,000, complicates the diagnostic process [4]. Some studies even suggest that the number of rare diseases could be as high as 10,000 [5].

### 1.2 Problem Statement

Despite advancements in medical science, the diagnostic process for rare diseases remains fraught with inefficiencies. More than 45% of patients with rare diseases remain undiagnosed or receive incorrect diagnoses [6], which can lead to inappropriate treatments and delayed access to potentially life-saving interventions. Misdiagnosis not only prolongs patient suffering but also places a significant burden on healthcare systems, with needless visits, tests, and treatments contributing to rising costs.

The urgency to enhance diagnostic accuracy and reduce time-to-diagnosis through innovative solutions is paramount. DxGPT, an experimental platform developed by Foundation 29, aims to address this critical need by exploring the use of large language models (LLMs) for rare disease diagnosis. By leveraging the vast knowledge and pattern recognition capabilities of LLMs, DxGPT seeks to assist healthcare professionals in navigating the complex landscape of rare diseases, potentially accelerating the diagnostic process and improving patient outcomes.

The platform’s goal is to provide a powerful tool that can generate accurate diagnostic hypotheses based on patient symptoms and clinical data, helping to guide physicians towards the correct diagnosis more efficiently. By harnessing the power of AI and natural language processing, DxGPT aims to complement human expertise, serving as a decision support system that can suggest potential rare diseases and provide relevant medical information to aid in the diagnostic process.

### 1.3 Objectives

This study aims to evaluate the effectiveness of DxGPT, a conversational assistant platform utilizing various large language models like GPT-4 [7], Claude3 [8], Gemini [9], and others, in generating accurate diagnostic suggestions. The objectives are to assess the diagnostic accuracy of DxGPT for synthetic and real-world patient descriptions representing cases of rare diseases, and to compare the performance across different AI models.

### 1.4 Significance

Large language models have demonstrated significant potential in various natural language processing tasks, including those relevant to medical diagnostics [10]. By leveraging state-of-the-art LLMs, DxGPT could transform medical practice by improving diagnostic accuracy, predicting disease progression, and supporting clinical decision-making [11]. This study’s results could lead to faster referrals to experts, better patient outcomes, and more informed clinical decision-making. Furthermore, the knowledge obtained from evaluating different models’ performance could inform future advances in AI-supported healthcare technologies.

## 2 Methodology

### 2.1 Data

The study utilizes several datasets:

- Synthetic Dataset: Created using GPT-4 (0613) with the prompt:

~~~
“Summarize in one paragraph the chief complaints
and notable findings that would be consistent with
early stages of {disease}, for a new patient coming to
primary care who has no clear diagnosis upon arrival.
Do not explicitly state {disease}.”
~~~

This dataset includes 200 synthetic patient cases derived from the top 200 rare diseases listed in Orphanet [12], featuring subtle symptom descriptions and additional noise.

- Real-World Data: Comprises two parts, leveraging the easily accessible datasets and benchmarks provided by Chen et al. in RareBench [13], which were instrumental to this paper:

1. 200 publicly available cases from RAMEDIS dataset, focusing on rare diseases with lists of Human Phenotype Ontology (HPO) symptoms.
2. 75 publicly available cases from Peking Union Medical College Hospital (PUMCH), which also consist of HPO symptom lists. Full dataset access has been requested and we are in discussions with the RareBench [13] researchers to further collaborate in this direction.

### 2.2 Evaluation Metrics

The evaluation is guided by two principal metrics:

- Strict Accuracy (P1): The rate at which the top diagnostic suggestion matched the actual diagnosis.
- Top-5 Accuracy (P1 + P5): The frequency of the correct diagnosis appearing within the top five suggestions.

### 2.3 Models Evaluated

The AI models selected for evaluation were chosen based on several criteria: they were highly ranked in public benchmarks, offered competitive pricing and latency, and had robust and easy API availability. This selection, while guided by these factors, also considered the practical constraints of our project timeline. The models evaluated include:

- Closed source/weights or proprietary models: GPT-4 0613 [7], GPT-4 Turbo 1106, GPT-4 Turbo 0409, Claude 3 Opus [8], Claude 3 Sonnet [8], Gemini 1.5 Pro [9].
- Open source/weights models: Llama 2 7B [14], Llama 3 8B, Llama 3 70B, Mistral 7B [15], Mixtral8x7B [16], Mixtral8x22B, Cohere Command R+.

Additional models may be included based on their emergence and relevance in the future.

### 2.4 Scripts and Methodology

An automated LLM pipeline was developed for this study, utilizing our standard diagnostic prompt from the web-based DxGPT platform. The evaluation script, which includes potential limitations of using LLMs like GPT-4 (0613) for automatic evaluation, will be detailed in the paper appendix and the related code is available at https://github.com/foundation29org/dxgpt_testing/. Alternatives for automatic evaluation will also be proposed and are being evaluated at the moment of publication.

## 3 Results

### 3.1 Diagnostic Accuracy Results

**Figure 1:**
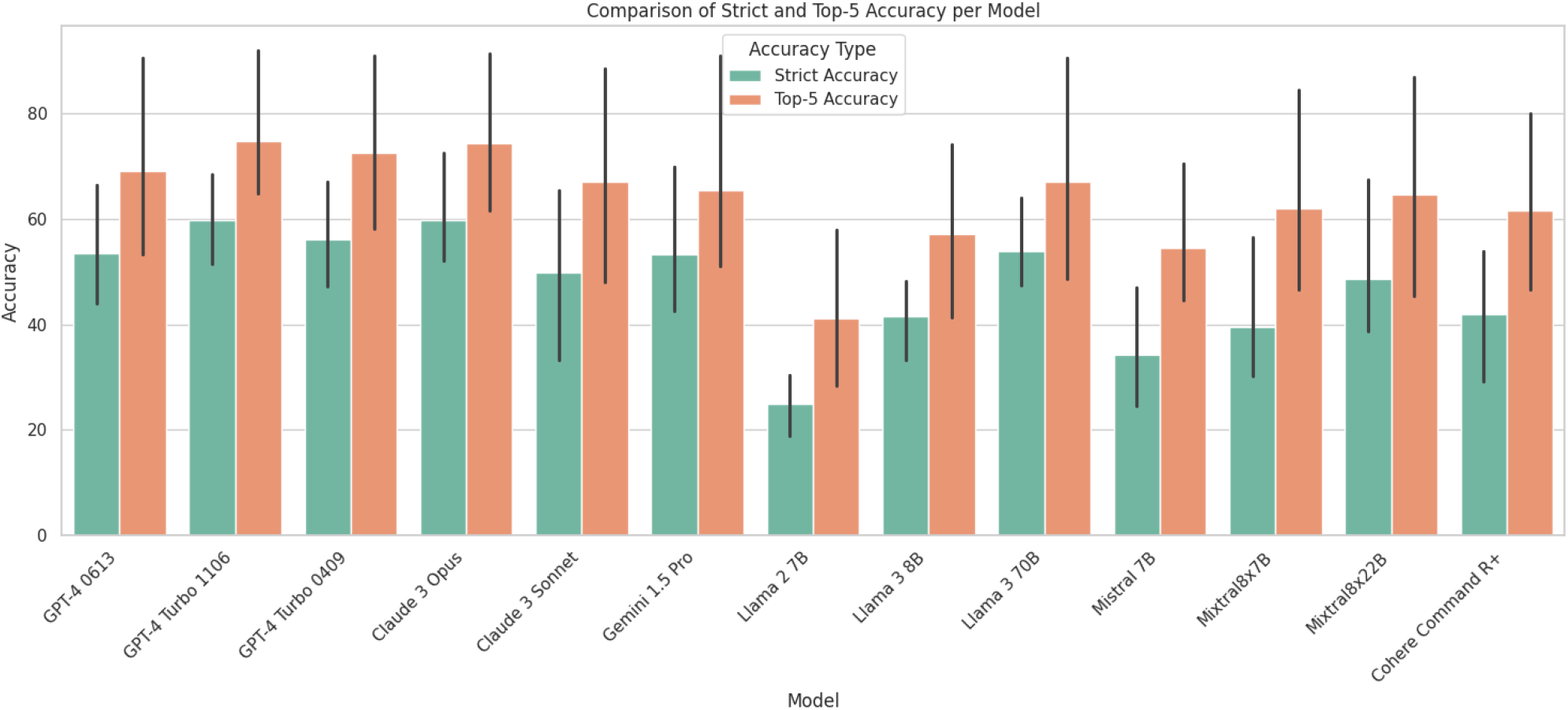
Comparative diagnostic accuracy of all the models on all datasets.

#### 3.1.1 Accuracy on Synthetic Dataset

**Table 1:** Diagnostic accuracy of closed models in our synthetic dataset (200 cases)

| Model | Strict Accuracy (P1) | Top-5 Accuracy (P1 + P5) |
| --- | --- | --- |
| GPT-4 0613 | 66.50% | 90.50% |
| GPT-4 Turbo 1106 | 68.50% | **92.00%** |
| GPT-4 Turbo 0409 | 67.00% | 91.00% |
| Claude 3 Opus | **72.50%** | 91.50% |
| Claude 3 Sonnet | 65.50% | 88.50% |
| Gemini 1.5 Pro | 70.00% | 91.00% |

**Table 2:** Diagnostic accuracy of open models in our synthetic dataset (200 cases)

| Model | Strict Accuracy (P1) | Top-5 Accuracy (P1 + P5) |
| --- | --- | --- |
| Llama 2 7B | 30.50% | 58.00% |
| Llama 3 8B | 48.22% | 74.11% |
| Llama 3 70B | 64.00% | **90.5%** |
| Mistral 7B | 47.00% | 70.50% |
| Mixtral8x7B | 56.50% | 84.50% |
| Mixtral8x22B | **67.50%** | 87.00% |
| Cohere Command R+ | 54.00% | 80.00% |

**Figure 2:**
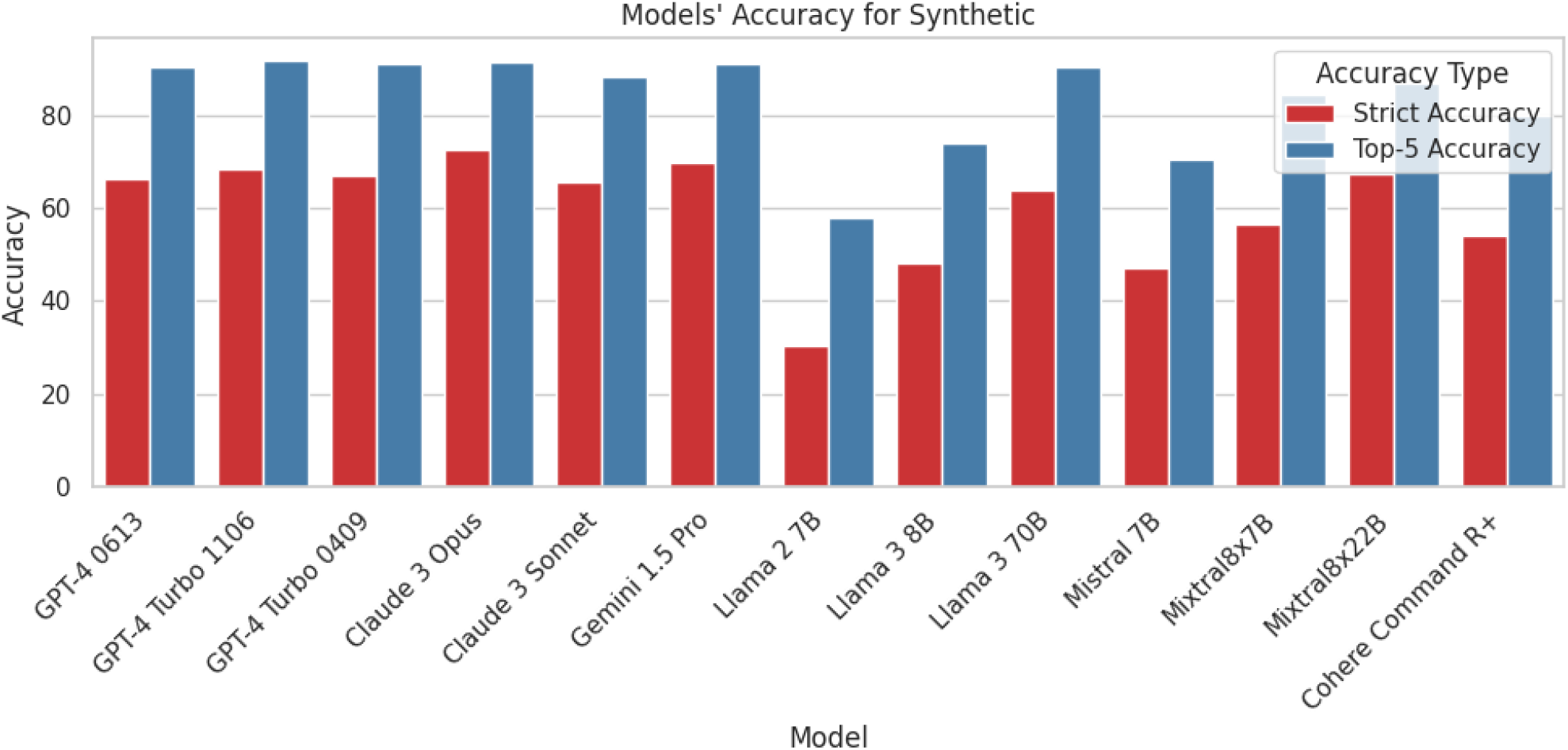
Comparative diagnostic accuracy of all the models on synthetic dataset.

#### 3.1.2 Accuracy on RAMEDIS Data

**Table 3:** Diagnostic accuracy of closed models in RAMEDIS data (200 cases)

| Model | Strict Accuracy (P1) | Top-5 Accuracy (P1 + P5) |
| --- | --- | --- |
| GPT-4 0613 | 50.25% | 63.32% |
| GPT-4 Turbo 1106 | 51.50% | 67.50% |
| GPT-4 Turbo 0409 | 54.04% | 68.69% |
| Claude 3 Opus | **55.00%** | **70.00%** |
| Claude 3 Sonnet | 50.50% | 64.50% |
| Gemini 1.5 Pro | 42.63% | 51.05% |

**Table 4:** Diagnostic accuracy of open models in RAMEDIS data (200 cases)

| Model | Strict Accuracy (P1) | Top-5 Accuracy (P1 + P5) |
| --- | --- | --- |
| Llama 2 7B | 25.63% | 37.19% |
| Llama 3 8B | 43.00% | 56.00% |
| Llama 3 70B | **53.54%** | **62.12%** |
| Mistral 7B | 24.62% | 48.24% |
| Mixtral8x7B | 31.82% | 55.05% |
| Mixtral8x22B | 39.59% | 61.93% |
| Cohere Command R+ | 29.29% | 58.08% |

**Figure 3:**
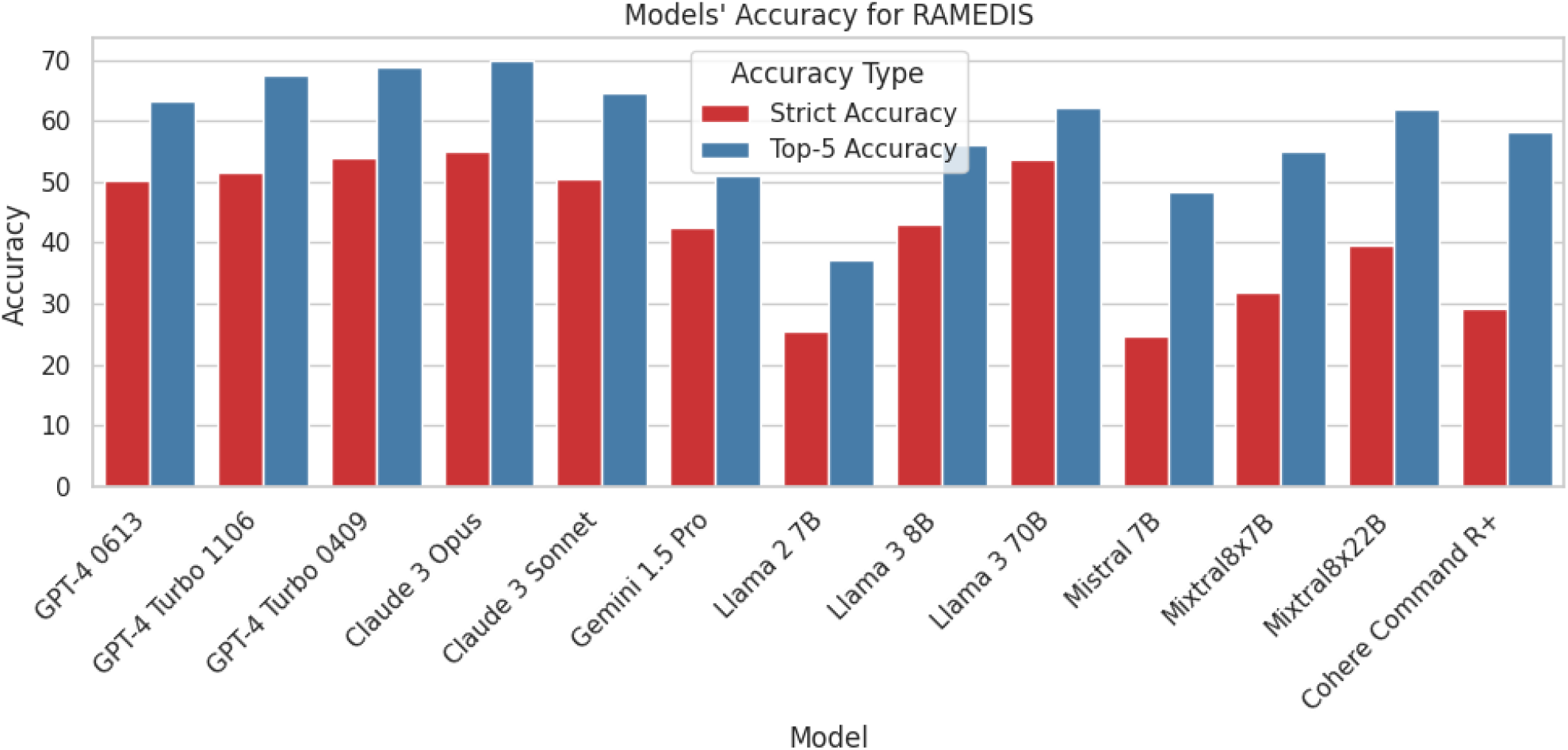
Comparative diagnostic accuracy of all the models on RAMEDIS dataset.

#### 3.1.3. Accuracy on Peking Union Medical College Hospital (PUMCH)

**Table 5:**
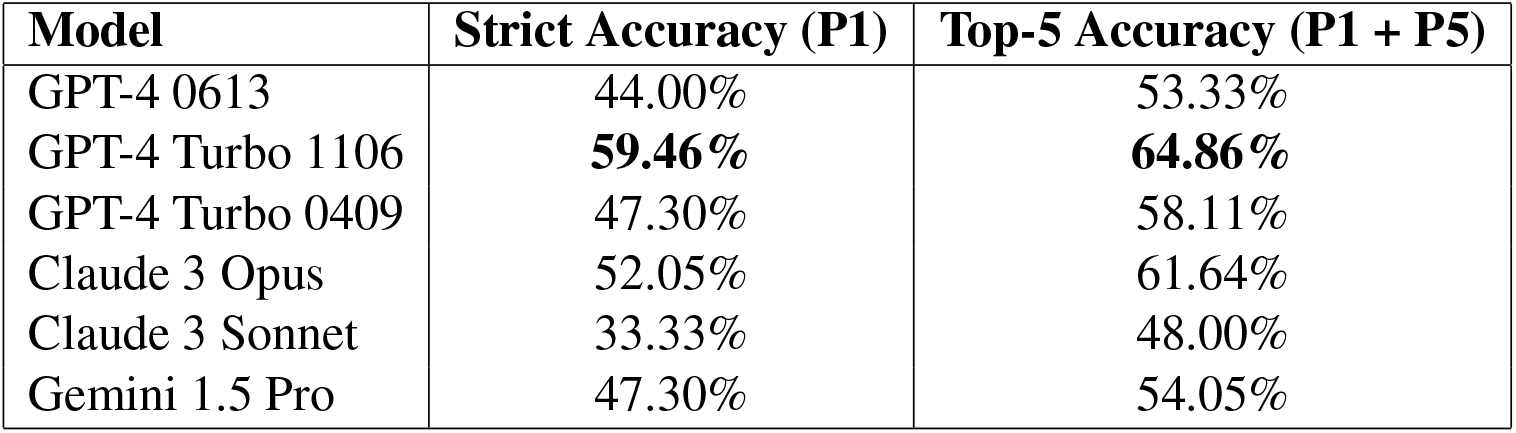
Diagnostic accuracy of closed models in PUMCH data (75 cases)

3.1.3 Accuracy on Peking Union Medical College Hospital (PUMCH)
| Model | Strict Accuracy (P1) | Top-5 Accuracy (P1 + P5) |
| --- | --- | --- |
| GPT-4 0613 | 44.00% | 53.33% |
| GPT-4 Turbo 1106 | **59.46%** | **64.86%** |
| GPT-4 Turbo 0409 | 47.30% | 58.11% |
| Claude 3 Opus | 52.05% | 61.64% |
| Claude 3 Sonnet | 33.33% | 48.00% |
| Gemini 1.5 Pro | 47.30% | 54.05% |

**Table 6:** Diagnostic accuracy of open models in PUMCH data (75 cases)

| Model | Strict Accuracy (P1) | Top-5 Accuracy (P1 + P5) |
| --- | --- | --- |
| Llama 2 7B | 18.92% | 28.38% |
| Llama 3 8B | 33.33% | 41.33% |
| Llama 3 70B | **44.44%** | **48.61%** |
| Mistral 7B | 31.08% | 44.59% |
| Mixtral8x7B | 30.14% | 46.58% |
| Mixtral8x22B | 38.67% | 45.33% |
| Cohere Command R+ | 42.67% | 46.67% |

**Figure 4:**
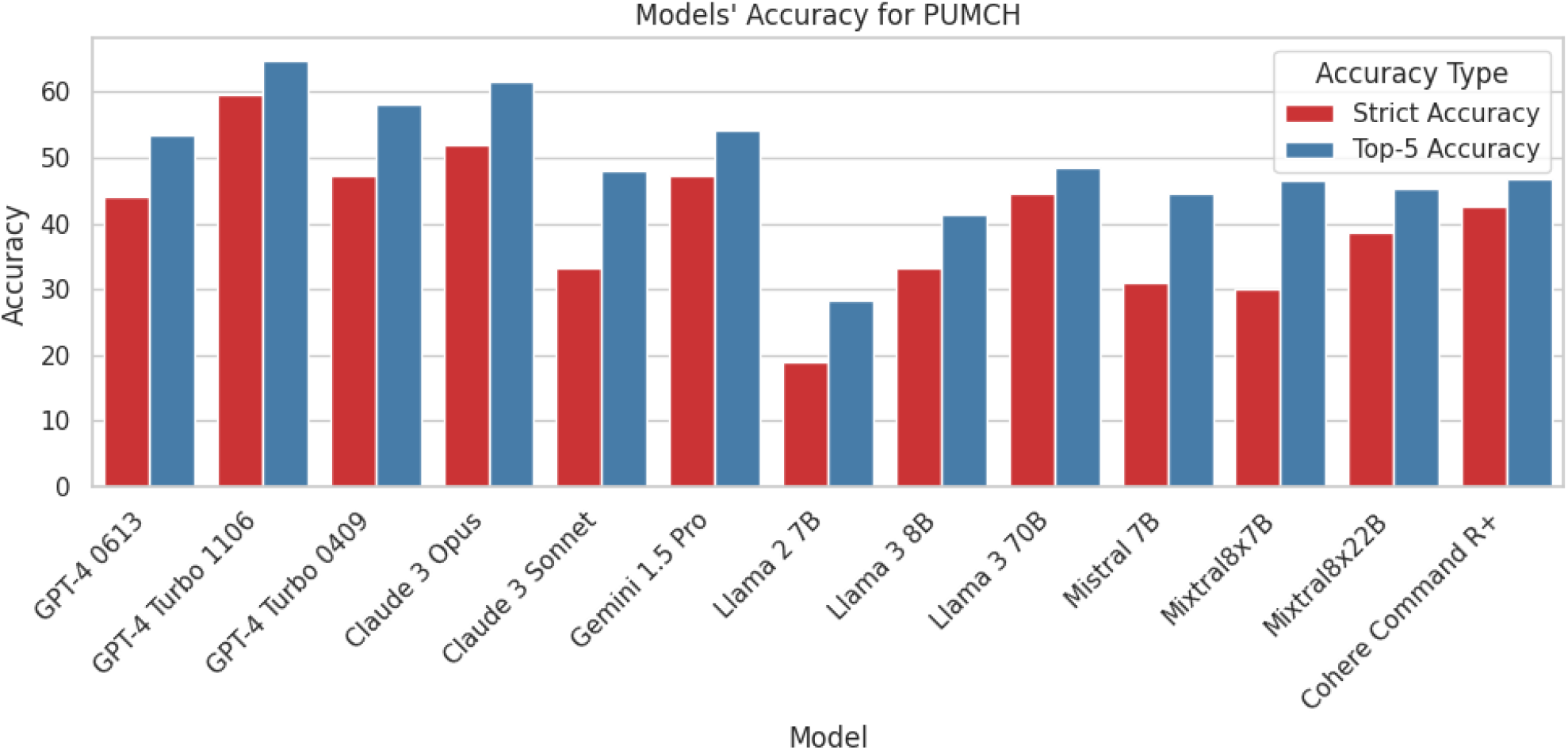
Comparative diagnostic accuracy of all the models on PUMCH dataset.

## 4 Discussion

The results reveal a nuanced landscape regarding the diagnostic accuracy of DxGPT and the various large language models evaluated. Several key observations can be made:

### 4.1 Synthetic Dataset Performance

- The closed models like GPT-4, Claude, and Gemini exhibited relatively high accuracy on the synthetic rare disease dataset, with Strict Accuracy ranging from 65.5% to 72.5% and Top-5 Accuracy from 88.5% to 92%.
- Open models like Llama, Mistral, Mixtral, and Cohere showed moderate to high accuracy, with Strict Accuracy ranging from 30.5% to 67.5% and Top-5 Accuracy from 58% to 90.5%.
- This suggests that current LLMs can effectively leverage the symptom descriptions in synthetic cases to generate accurate diagnostic suggestions for rare diseases.

### 4.2 Real-World Dataset Performance

- Accuracy dropped notably on real-world datasets like RAMEDIS and PUMCH compared to synthetic cases, as expected.
- On RAMEDIS rare disease cases, Claude 3 Opus model demonstrated 55% Strict Accuracy and 70% Top-5 Accuracy, outperforming other closed models which followed with Strict Accuracy in the range of 50% to 54% and Top-5 Accuracy in the range of 63% to 68.6%. Gemini was the worst closed model with 42% Strict Accuracy and 51% Top-5 Accuracy. Open models like Llama 3 and Mixtral showed moderate accuracy.
- The PUMCH dataset proved challenging for all models, with the highest Strict Accuracy at 59.46% (GPT-4 Turbo 1106) and Top-5 Accuracy at 64.86%. Llama 3 70B was the best open model with 44.44% Strict Accuracy and 48.61% Top-5 Accuracy.

### 4.3 Variability Across Models

- While GPT-4 variants consistently ranked among the top performers, there was some variability in accuracy across different model versions. For example, GPT-4 Turbo 1106 outperformed GPT-4 Turbo 0409 on the PUMCH dataset (59.46% vs 47.30% Strict Accuracy), while the reverse was true on the RAMEDIS data (51.5% vs 54.04% Strict Accuracy). This suggests that even within the same model family, specific versions may have strengths or weaknesses for certain types of clinical data.
- The open models exhibited an even wider range of performance. Llama 3 70B stood out as a strong performer, achieving the highest accuracy among open models on all three datasets. It even rivaled some of the closed models, especially on the synthetic data (64% Strict Accuracy, 90.5% Top-5 Accuracy).
- In contrast, models like Llama 2 7B and Mistral 7B struggled more, particularly on the real-world RAMEDIS and PUMCH datasets where their Strict Accuracy fell below 32%. This gap between the best and worst performing open models highlights the importance of model architecture, training data, and other factors in determining diagnostic capabilities.
- Interestingly, some of the larger open models like Mixtral8x22B showed promise, outperforming their smaller counterparts. On the synthetic data, Mixtral8x22B achieved 67.5% Strict Accuracy, on par with some of the closed models. However, this advantage diminished on the real-world datasets, underscoring the challenges in translating performance to clinical practice.

### 4.4 Strengths and Limitations

#### 4.4.1 Strengths

The study leverages a diverse dataset, including synthetic and real-world data from RAMEDIS and PUMCH, enhancing the generalizability and robustness of the findings. The inclusion of multiple AI models, both closed and open source, provides a broad perspective on AI capabilities in diagnostic processes. Evaluation metrics such as Strict Accuracy (P1) and Top-5 Accuracy (P1 + P5) are clinically relevant, offering insights into the models’ practical utility in healthcare. The detailed methodology, including data preprocessing, model inference, and evaluation processes, is thoroughly documented in the appendix, enhancing the study’s reproducibility and transparency.

#### 4.4.2 Limitations

Despite its strengths, the study faces several limitations. The lack of direct clinical validation means the practical utility of the AI models in real-world clinical settings remains untested. The synthetic dataset, generated using GPT-4, may carry biases inherent to the model, and the real-world datasets may not fully represent the diversity of rare diseases. Furthermore, while we have included code for calculating p-values in the accompanying repository, we have chosen not to discuss these statistical details in the main text of the paper, as they do not align with the primary objectives of this study.

The study does not delve into the interpretability of the AI models, which is crucial for their acceptance and trust among healthcare professionals. Additionally, there is no direct comparison with the diagnostic accuracy of human experts, which is necessary to fully assess the AI models’ effectiveness. Lastly, the use of LLMs as automatic evaluators for batch examining the results against ground truths presents its own set of challenges as stated in Shankar et al. [17]. This method, while efficient, might overestimate the models’ performance suggesting a need for validation through human expert evaluation in future studies.

## 5 Conclusion and Future Work

This comprehensive evaluation of DxGPT and various large language models has yielded valuable insights into their diagnostic capabilities for rare diseases. The findings underscore the potential of LLMs to enhance clinical reasoning and support accurate diagnosis, particularly when leveraging detailed symptom data.

However, the study also reveals challenges in translating synthetic performance to real-world clinical scenarios, where factors like data quality, disease complexity, and model limitations come into play. While closed models like GPT-4 demonstrated promising accuracy, there is still room for improvement, especially on rare disease cases and datasets with limited information.

Looking ahead, continued research and development efforts are crucial to refine these AI-driven diagnostic tools. Potential avenues include:

1. Expanding and diversifying training data to improve model performance on real-world cases.
2. Integrating multimodal data sources, such as imaging [18, 19] and electronic health records [20, 21], to enhance the robustness and applicability of models in clinical settings.
3. Exploring advanced techniques like many-shot in-context learning [22], sophisticated prompt engineering [23, 24], and multi-task fine-tuning to boost model adaptability and accuracy.
4. Developing hybrid approaches that combine LLMs with other AI techniques, such as knowledge graphs [25] and reasoning engines [26], to leverage complementary strengths and multimodal capabilities.
5. Conducting prospective clinical studies to validate the real-world impact of AI-assisted diagnosis on patient outcomes and healthcare costs.
6. Additionally, enhancing interpretability of AI models [27] and their integration into clinical workflows to facilitate continuous learning from clinician feedback, thereby improving the models’ accuracy and relevance.

Ultimately, the successful integration of AI into medical diagnostics hinges on a collaborative effort between researchers, clinicians, and regulatory bodies to ensure safety, efficacy, and ethical deployment of these powerful technologies.

### 5.1 Ongoing Clinical Studies

We have initiated real-world clinical studies involving DxGPT with real doctors and human evaluators in different hospitals and healthcare systems. These studies are designed to further validate the findings of this research and enhance the diagnostic capabilities of DxGPT. By integrating feedback from healthcare professionals directly involved in patient care, we aim to refine the model’s accuracy and utility in practical settings.

### 5.2 Ethical and Regulatory Considerations

Addressing ethical considerations, such as data security, patient privacy, and algorithmic bias, is crucial. Collaborative efforts with regulatory bodies will be essential to ensure that AI diagnostic tools adhere to clinical and ethical standards before widespread implementation.

It is important to emphasize that the primary goal of AI diagnostic tools like DxGPT is to assist patients by improving the accuracy and speed of diagnosis, particularly for rare diseases and complex cases that often face significant unmet medical needs. By leveraging the power of large language models and vast medical knowledge bases, AI tools have the potential to greatly reduce diagnostic delays and improve patient outcomes.

However, the evaluation of AI diagnostic accuracy should be approached with a balanced perspective. While rigorous testing and validation are essential, it is important to acknowledge that the current diagnostic accuracy of human physicians is not always well-established or consistently measured. Therefore, setting excessively high accuracy thresholds for AI tools without considering the real-world performance of human diagnosticians may create an unfair double standard.

DxGPT and similar AI diagnostic tools should be viewed as proof-of-concept systems that demonstrate the potential of this technology to augment and support human medical expertise. They are not intended to replace physicians but rather to serve as powerful tools that can help generate accurate diagnostic hypotheses and guide clinical decision-making. As such, the accuracy expectations for these AI tools should be benchmarked against the performance of human doctors, taking into account the inherent challenges and uncertainties in medical diagnosis.

Moreover, AI diagnostic tools offer unique advantages that can complement human skills. They can rapidly process vast amounts of medical literature, identify subtle patterns and associations, and consider rare diseases that may be overlooked by physicians.

As we continue to develop and refine AI diagnostic tools, it is essential to maintain an open and transparent dialogue among researchers, clinicians, patients, and regulatory bodies. Engaging all stakeholders in the process will help ensure that these technologies are developed and implemented in an ethical, responsible, and patient-centric manner. Regular audits and assessments should be conducted to monitor the performance and fairness of AI models, and mechanisms should be in place to allow for continuous improvement based on real-world feedback and outcomes.

In conclusion, while the development of AI diagnostic tools like DxGPT raises important ethical considerations, these should not overshadow the immense potential of this technology to address unmet medical needs and improve patient care. By setting realistic accuracy expectations, fostering collaboration between AI and human experts, and ensuring robust ethical and regulatory oversight, we can harness the power of AI to revolutionize medical diagnosis and ultimately benefit patients worldwide. DxGPT and similar proof-of-concept systems represent an exciting step forward in this journey, paving the way for a future where AI and human intelligence work together to provide faster, more accurate, and more equitable healthcare for all.

## Data Availability

All data produced are available online at:
https://huggingface.co/datasets/chenxz/RareBench
and
https://github.com/foundation29org/dxgpt_testing

https://github.com/foundation29org/dxgpt_testing

## 6 Appendices

Here we provide additional details on the methodology, scripts, and prompts used in the evaluation of DxGPT and large language models.

As mentioned in the paper, the evaluation script for DxGPT and LLMs is available at https://github.com/foundation29org/dxgpt_testing/. The script includes the following components:

- Synthetic dataset generation: Using GPT-4 to create 200 synthetic patient cases with subtle symptom descriptions for rare diseases.
- Data preprocessing: Loading and formatting the synthetic and real-world datasets for input to the LLMs.
- Model inference: Running the LLMs on the patient cases to generate diagnostic suggestions.
- Automatic evaluation metrics: Calculating the Strict Accuracy and Top-5 Accuracy based on the model outputs and ground truth diagnoses.
- Results analysis: Aggregating and visualizing the diagnostic accuracy results for comparison across models and datasets.

Here we also provide the detailed prompts used for generating synthetic patient cases, diagnosing them, and evaluating the model outputs against the ground truth diagnoses. The prompts are designed to simulate a clinical scenario where a doctor interacts with a conversational AI tool to diagnose rare diseases based on patient descriptions.

Synthetic patient case prompt:

~~~
     “Summarize in one paragraph the chief complaints
     and notable findings that would be consistent with
     early stages of {disease}, for a new patient coming to
     primary care who has no clear diagnosis upon arrival.
     Do not explicitly state {disease}.”
~~~

Diagnostic prompt:

~~~
     “Behave like a hypothetical doctor who has to do
     a diagnosis for a patient. Give me a list of potential
     diseases with a short description. Shows for each
     potential diseases always with ‘\n\n+’ and a number,
     starting with ‘\n\n+1’, for example ‘\n\n+23.’ (never
     return ‘\n\n-’), the name of the disease and finish
     with ‘:’. Dont return ‘\n\n-’, return ‘\n\n+’ instead.
     You have to indicate which symptoms the patient
     has in common with the proposed disease and which
     symptoms the patient does not have in common. The
     text is ‘\n\n Symptoms:{description}‘“
~~~

Evaluation prompt:

~~~
     “Behave like a medical doctor reviewing patient diagnoses.
     You will be given a Ground Truth diagnosis (GT) and 5
     Predicted diagnoses (P1-P5).
     Compare the GT to the predictions and return a
     classification:
     If GT exactly matches P1, return “P1”.
     If GT is contained within or is a broader term for P1-P5,
     return “P5”.
     If GT does not match any of P1-P5, return “P0”.
     The GT may be a more general diagnosis, while predictions
     may include specific conditions.
     Broadly match GT to any prediction it reasonably
     encompasses.
     ----------------------------------------
     The text is: GT: {gt} Predictions:
     {predictions}
     ----------------------------------------
     Return either “P1”, “P5”, or “P0”. Do not return
     any other text.”
~~~

## References

1. Benito-Lozano, J., Lopez-Villalba, B., Arias-Merino, G., Posada de la Paz, M. & Alonso-Ferreira, V. Diagnostic delay in rare diseases: data from the Spanish rare diseases patient registry. Orphanet Journal of Rare Diseases 17, 418. ISSN: 1750-1172. 10.1186/s13023-022-02530-3 (Nov. 2022).

2. Marwaha, S., Knowles, J. W. & Ashley, E. A. A guide for the diagnosis of rare and undiagnosed disease: beyond the exome. Genome Medicine 14. ISSN: 1756-994X. 10.1186/s13073-022-01026-w (Feb. 2022).

3. Ronicke, S. et al. Can a decision support system accelerate rare disease diagnosis? Evaluating the potential impact of Ada DX in a retrospective study. Orphanet Journal of Rare Diseases 14. ISSN: 1750-1172. 10.1186/s13023-019-1040-6 (Mar. 2019).

4. Haendel, M. et al. How many rare diseases are there? Nature Reviews Drug Discovery 19, 77–78. ISSN: 1474-1784. 10.1038/d41573-019-00180-y (Nov. 2019).

5. Hartley, T. et al. The unsolved rare genetic disease atlas? An analysis of the unexplained phenotypic descriptions in OMIM®. American Journal of Medical Genetics Part C: Seminars in Medical Genetics 178, 458–463. eprint: https://onlinelibrary.wiley.com/doi/pdf/10.1002/ajmg.c.31662. https://onlinelibrary.wiley.com/doi/abs/10.1002/ajmg.c.31662 (2018).

6. Molster, C. et al. Survey of healthcare experiences of Australian adults living with rare diseases. Orphanet Journal of Rare Diseases 11. ISSN: 1750-1172. 10.1186/s13023-016-0409-z (Mar. 2016).

7. OpenAI et al. GPT-4 Technical Report. arXiv: 2303.08774 [cs.CL] (2024).

8. Anthropic. The Claude 3 Model Family: Opus, Sonnet, Haiku. Anthropic. https://www-cdn.anthropic.com/de8ba9b01c9ab7cbabf5c33b80b7bbc618857627/Model_CardClaude_3.pdf (2024).

9. Team, G. et al. Gemini: A Family of Highly Capable Multimodal Models. arXiv: 2312.11805 [cs.CL] (2024).

10. Karabacak, M. & Margetis, K. Embracing Large Language Models for Medical Applications: Opportunities and Challenges. Cureus. ISSN: 2168-8184. 10.7759/cureus.39305 (May 2023).

11. Nori, H., King, N., McKinney, S. M., Carignan, D. & Horvitz, E. Capabilities of GPT-4 on Medical Challenge Problems. https://arxiv.org/abs/2303.13375 (2023).

12. Prevalence of rare diseases by alphabetical list https://www.orpha.net/pdfs/orphacom/cahiers/docs/GB/Prevalence_of_rare_diseases_by_alphabetical_list.pdf. Accessed: 2024-01-29.

13. Chen, X. et al. RareBench: Can LLMs Serve as Rare Diseases Specialists? arXiv: 2402.06341 [cs.CL] (2024).

14. Touvron, H. et al. Llama 2: Open Foundation and Fine-Tuned Chat Models. arXiv: 2307.09288 [cs.CL] (2023).

15. Jiang, A. Q. et al. Mistral 7B. arXiv: 2310.06825 [cs.CL] (2023).

16. Jiang, A. Q. et al. Mixtral of Experts. arXiv: 2401.04088 [cs.LG] (2024).

17. Shankar, S., Zamfirescu-Pereira, J. D., Hartmann, B., Parameswaran, A. G. & Arawjo, I. Who Validates the Validators? Aligning LLM-Assisted Evaluation of LLM Outputs with Human Preferences. arXiv: 2404.12272 [cs.HC] (2024).

18. Saab, K. et al. Capabilities of Gemini Models in Medicine. arXiv: 2404.18416 [cs.AI] (2024).

19. Yang, L. et al. Advancing Multimodal Medical Capabilities of Gemini. arXiv: 2405.03162 [cs.CV] (2024).

20. Hsieh, T.-C. et al. GestaltMatcher facilitates rare disease matching using facial phenotype descriptors. Nature Genetics 54, 349–357. ISSN: 1546-1718. 10.1038/s41588-021-01010-x (Feb. 2022).

21. Kurokawa, R. et al. Diagnostic Performance of Claude 3 from Patient History and Key Images in Diagnosis Please Cases. medRxiv. eprint: https://www.medrxiv.org/content/early/2024/04/14/2024.04.11.24305622.full.pdf. https://www.medrxiv.org/content/early/2024/04/14/2024.04.11.24305622 (2024).

22. Agarwal, R. et al. Many-Shot In-Context Learning. arXiv: 2404.11018 [cs.LG] (2024).

23. Abdullahi, T., Singh, R. & Eickhoff, C. Learning to Make Rare and Complex Diagnoses With Generative AI Assistance: Qualitative Study of Popular Large Language Models. JMIR Med Educ 10, e51391. ISSN: 2369-3762. http://www.ncbi.nlm.nih.gov/pubmed/38349725 (Feb. 2024).

24. Oniani, D. et al. Large Language Models Vote: Prompting for Rare Disease Identification. arXiv: 2308.12890 [cs.CL] (2024).

25. Chandak, P., Huang, K. & Zitnik, M. Building a knowledge graph to enable precision medicine. Scientific Data 10. ISSN: 2052-4463. 10.1038/s41597-023-01960-3 (Feb. 2023).

26. Tu, T. et al. Towards Conversational Diagnostic AI. arXiv: 2401.05654 [cs.AI] (2024).

27. Bricken, T. et al. Towards Monosemanticity: Decomposing Language Models With Dictionary Learning. Transformer Circuits Thread. https://transformer-circuits.pub/2023/monosemantic-features/index.html (2023).

